# Genomics of Aminoglycoside Resistance in *Pseudomonas aeruginosa* Bloodstream Infections at a United States Academic Hospital

**DOI:** 10.1101/2021.01.15.21249897

**Authors:** Giancarlo Atassi, Marc Scheetz, Sophia Nozick, Nathaniel J. Rhodes, Megan Murphy-Belcaster, Katherine R Murphy, Egon A. Ozer, Alan R. Hauser

## Abstract

*Pseudomonas aeruginosa* is a frequent cause of antibiotic-resistant infections. Although *P. aeruginosa* is intrinsically resistant to many antimicrobial agents, aminoglycosides are active against this organism in the absence of acquired resistance determinants and mutations. However, genes encoding aminoglycoside modifying enzymes (AMEs) are found in many strains that are resistant to these agents. We examined the prevalence of phenotypic resistance to the commonly used aminoglycosides gentamicin, tobramycin, and amikacin in a collection of 227 *P. aeruginosa* bloodstream isolates collected over two decades from a single U.S. academic medical institution. Resistance to these antibiotics was relatively stable over this time period. High-risk clones ST111 and ST298 were initially common but decreased in frequency over time. Whole genome sequencing identified relatively few AME genes in this collection compared to the published literature; only 14% of isolates contained an AME gene other than the ubiquitous *aph(3’)-IIb*. Of those present, only *ant(2”)-Ia* was associated with phenotypic resistance to gentamicin or to tobramycin. One extensively drug-resistant strain, PS1871, contained 5 AME genes, most of which were part of clusters of antibiotic resistance genes embedded within transposable elements. These findings suggest that AME genes play a relatively minor role in aminoglycoside resistance at our institution but that multidrug-resistant strains remain a problem.

## Introduction

*Pseudomonas aeruginosa* is a high-priority pathogen with a propensity for causing devastating opportunistic and nosocomial infections [1]. Complicating treatment of infections caused by this bacterium is significant intrinsic antimicrobial resistance (AMR) and a remarkable ability to mutate and acquire resistance determinants. As a result, many *P. aeruginosa* isolates are highly resistant to antibiotics, and a single strain may carry numerous antibiotic resistance genes [1-3]. Several of these multidrug resistant (MDR) and extensively drug-resistant (XDR) strains have disseminated globally and are referred to as “high-risk clones.” Four major sequence types of these high-risk clones have been characterized over the past decade: ST111, ST175, ST235, and ST298 [1][4]. High-risk clones account for substantial proportions of multidrug-resistant *P. aeruginosa* isolates in some parts of the world and are a major healthcare threat.

Two approaches have been taken to address the challenge of antibiotic-resistant organisms such as *P. aeruginosa*. At the clinical level, antimicrobial stewardship programs are now required at medical institutions. Their role is to promote judicious and appropriate use of antimicrobial agents through evidence-based practice, and they have generally provided cost savings and decreased inappropriate antibiotic use and nosocomial infections by resistant pathogens without leading to increased mortality [5-8]. At the diagnostic level, molecular identification of resistance genes and alleles via nucleic-acid amplification or whole genome sequencing is being pursued to rapidly identify resistant and susceptible organisms and thus allow more focused antimicrobial therapy [9]. However, genomic resistance determinants can only provide surrogate markers of AMR and must be correlated to clinical phenotypes to validate their utility.

AMR genotype/phenotype correlation studies with *P. aeruginosa* have been challenging [10-12]. In particular, efforts directed at predicting aminoglycoside resistance have met with mixed success. *P. aeruginosa* deploys an array of resistance mechanisms against this class of antibiotics. Isolates can inactivate aminoglycosides via aminoglycoside modifying enzymes (AMEs), of which there are three primary families: phosphorylators (aph), acetylators (AAC), and adenylators (ant) [13, 14]. Adding to this complexity is that individual AMEs have variable activities against the different aminoglycosides. *AAC(3), AAC(6’)-I, AAC(6’)-II*, and *ant(2”)* inactivate gentamicin; *AAC(3)-II, ant(2”), ant(4’)-I*, and *ant(4’)-II* inactivate tobramycin; *AAC(6’)-Ib, AAC(3)-X, ant(4’)-I, ant(4’)-II, aph(3’)-IIIa*, and *aph(3’)-VIb* inactivate amikacin. Other mechanisms of resistance include overexpression of efflux pumps (particularly the MexXY-OprM complex), modification of 16S rRNA molecules by methylases such as *rmtA* and *rmtB* (rendering aminoglycosides unable to effectively bind ribosomes), and decreased permeability to aminoglycosides [14].

Here, we examined aminoglycoside resistance in *P. aeruginosa* bloodstream isolates at Northwestern Memorial Hospital (NMH) in Chicago, U.S. We examined changes in the prevalence of aminoglycoside resistance and the presence of AME genes over two decades and whether these genes predicted aminoglycoside resistance in these isolates. Finally, we determined the complete whole genome sequence of a MDR isolate carrying four distinct AME genes to examine the context of these resistance genes.

## Materials and Methods

### Antimicrobial Susceptibility Testing

We obtained *P. aeruginosa* bloodstream isolates from three archived NMH collections cultured from 1999-2002, 2003-2009, and 2017-2019. All isolates were collected in accordance with and approval by the Northwestern University Institutional Review Board. Susceptibility testing was performed by broth microdilution for eight antibiotics (cefepime, ciprofloxacin, piperacillin-tazobactam, meropenem, colistin, amikacin, gentamicin, and tobramycin) as previously described [15], and susceptibility vs. nonsusceptibility (defined as resistant and intermediate classification) was determined using Clinical and Laboratory Standards Institute (CLSI) breakpoints [16]. For isolates not tested by broth microdilution, susceptibility results were generated by the NMH Clinical Microbiology Laboratory using the Vitek 2 platform (bioMérieux, Marcy l’Etoile, France).

### Whole genome sequencing

The genome sequences of the 1999-2002 isolates were previously published [17][4, 18]. For the remaining isolates, bacteria were grown and underwent DNA extraction as previously described [17]. Library preparations were performed using the Nextera XT DNA Prep Kit (Illumina, Inc., San Diego, CA, USA) following the manufacturer’s protocol. Short-read whole-genome sequencing was performed for all isolates using either the Illumina HiSeq or MiSeq platform to generate paired-end reads. Reads were trimmed using Trimmomatic (v0.36). Draft genomes were assembled from trimmed paired-end reads using SPAdes (v3.9.1). Draft genomes were further filtered to remove contigs shorter than 200□bp, with less than 5-fold mean read coverage, or with alignment to phiX. For PS1871, long-read sequencing was performed using the Nanopore MinION platform, and hybrid assembly with short-read Illumina sequences were performed as previously described [19]. Briefly, long-read sequencing libraries were prepared from unsheared genomic DNA using ligation sequencing kit SQK-LSK109 (Oxford Nanopore, UK) and sequenced on the MinION platform using a FLO-MIN106 flow cell. Guppy v3.4.5 was used to base call reads with the R9.4.1 high-accuracy model and to perform read quality filtering based on Q scores, demultiplexing, and barcode trimming. Assembly of nanopore reads was performed using Flye v2.8.1 to generate a single circularized contig [20]. Illumina reads were aligned to the assembly using BWA v0.7.17 [21], and assembly errors were corrected using Pilon v1.23 [22] with a minimum depth setting of 0.1. Serial read alignment and Pilon correction were performed sequentially until no further assembly corrections were generated. Custom software (Pilon Tools v0.1; https://github.com/egonozer/pilon_tools) was used to identify, manually assess, and correct any residual homopolymer assembly errors. Accession numbers for all newly sequenced isolates are pending.

### Detection of antibiotic resistance genes

Assembled libraries were submitted to the AMRFinderPlus tool (version 3.9.3, last accessed November 23, 2020) for detection of antimicrobial resistance genes and resistance-associated point mutations (Feldgarden et al., 2019) [23]. AMRFinderPlus is available from the National Center for Biotechnology Information (NCBI) at https://www.ncbi.nlm.nih.gov/pathogens/antimicrobial-resistance/AMRFinder/. The DDBJ Fast Annotation & Submission Tool (DFAST; v.1.2.4, last accessed January 3^rd^, 2021) was used to annotate specific genomic features of the PS1871 chromosome [24]. SnapGene Viewer (from Insightful Science; available at snapgene.com) was used to generate linear chromosomal maps of the local regions of PS1871 surrounding AME genes found with AMRFinder, and the results from DFAST were used to annotate surrounding features.

### Determination of MLST genotype

Each isolate’s MLST was determined using ResFinder (v4.1, last accessed November 23, 2020; Center for Genomic Epidemiology) [25, 26]. ST111, ST175, ST235, and ST298 isolates were designated high-risk clones [1, 4]. MLSTs were assigned by ResFinder using PubMLST, an open-access curated database [27] that relies on a previously described MLST scheme for *P. aeruginosa* utilizing the seven housekeeping genes *acsA, aroE, guaA, mutL, mutD, ppsA*, and *trpE* [28].

### Statistical Analysis

Changes in antibiotic susceptibility rates over time and changes in the number of isolates with at least one AME gene across each of the three time periods were analyzed using Chi-Square tests of linear trend to determine whether there was a significant trend across the different time periods. All Chi-Square tests of trend were performed in EpiInfo 7.2.4.0 (Centers for Disease Control & Prevention, Atlanta, GA). The remainder of the statistics were performed in Stata IC 16.1 (Statacorp LLC, College Station, TX). For phenotypic aminoglycoside resistance predictions based on the presence of single AME genes, logistic regressions were performed using the presence of each individual AME gene. The presence of *at least one* member of gene groups were also considered; AME genes were grouped based on their activities against a specific aminoglycoside. A gentamicin group was comprised of the following AME genes: *AAC(3), AAC(6)-I, AAC(6)-II*, or *ant(2”)*. Initially, a tobramycin group comprised of *ant(2”), AAC(3), ant(4)-I*, or *ant(4)-II* was also planned. However, only one of these tobramycin-active genes (*ant(2”)-Ia)* was found in our collection, so we did not perform this analysis. Logistic regression was completed with the gentamicin-active gene group for predicting categorical resistance and was performed in stepwise fashion with P<0.2 used for removal from the model. Results are reported from the final model in the form of odds ratios with confidence intervals and p-values.

## Results

### Changes in Aminoglycoside Resistance of Pseudomonas aeruginosa Over Time at NMH

We examined aminoglycoside resistance in a total of 227 archived *P. aeruginosa* bloodstream isolates from three time periods: 93 from 1999-2002, 101 from 2003-2009, and 33 from 2017-2019. Susceptibility rates to gentamicin, tobramycin, and amikacin were measured.

Susceptibility rates to amikacin were highest and remained stable across these time periods, ranging from 92-94% (Figure 1). Susceptibility rates to tobramycin were somewhat lower (85-87%) but also remained stable. In contrast, susceptibility rates to gentamicin were lower still and more variable. They were 73% in 1999-2002, 87% in 2003-2009, and then 76% in 2017-2019 (Chi-Square test of linear trend, χ = 1.971, p = 0.160). We compared these susceptibility rates to those of five other commonly used antipseudomonal antibiotics: piperacillin-tazobactam, cefepime, meropenem, ciprofloxacin, and colistin. For each of these except colistin, susceptibility rates had not undergone a statistically significant change across the three time periods (Figure 1). Susceptibility rates were lowest for ciprofloxacin, for which only 57-70% of isolates were susceptible. For colistin, susceptibility rates were 92% in the first cohort, decreased to 69% in the second cohort, and then increased to 82% in the third cohort (Chi-Square test of linear trend, χ = 11.799, p < 0.001). These results suggest that *P. aeruginosa* isolates at NMH have remained relatively susceptible to amikacin and tobramycin but that susceptibility rates to gentamicin have been more variable.

**Figure 1.**
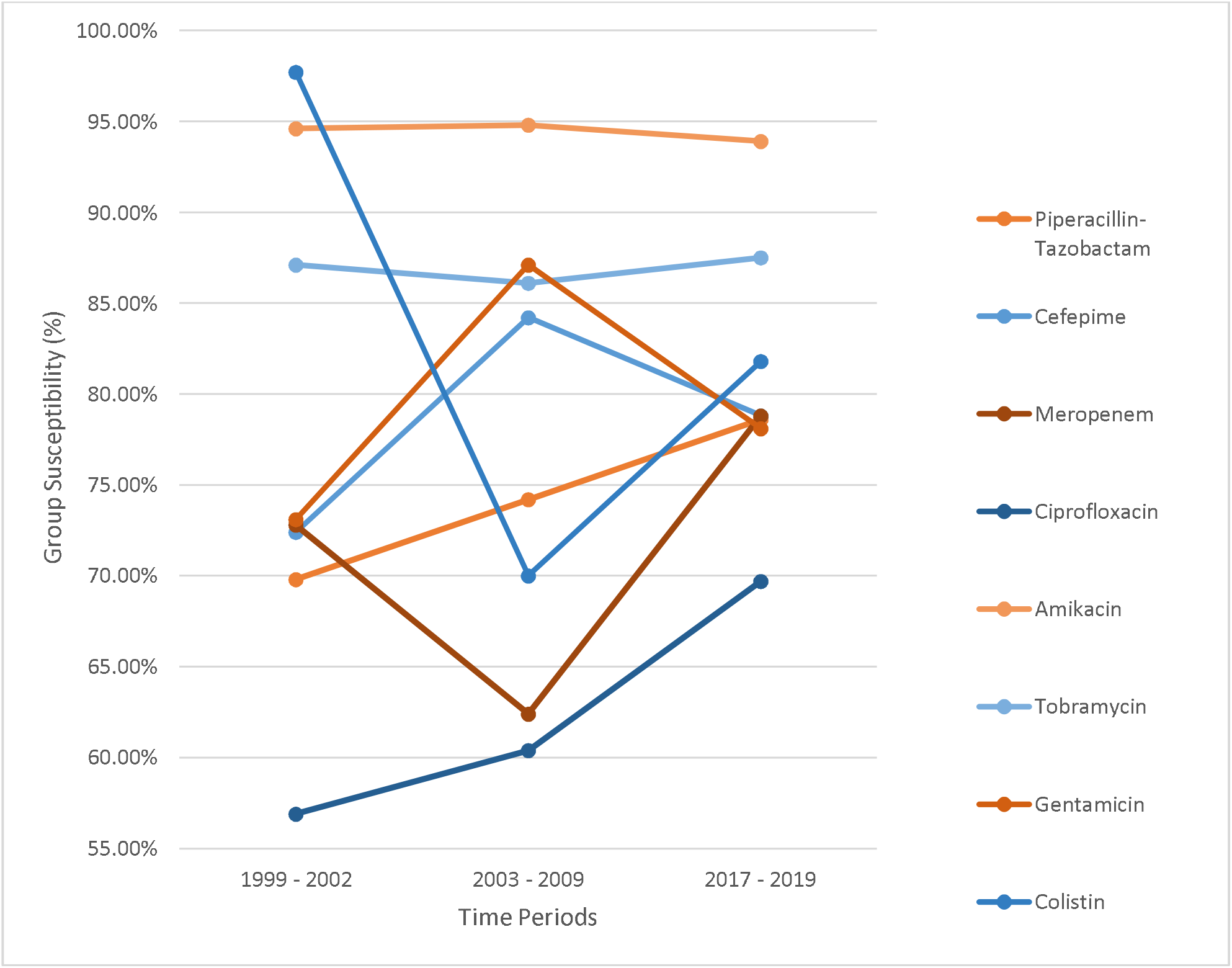
Susceptibility rates of *P. aeruginosa* isolates by antibiotic and time period.

### Prevalence and Changes in Multilocus Sequence Types over Time at NMH

High-risk clones are responsible for a substantial proportion of antibiotic resistance, including aminoglycoside resistance, in *P. aeruginosa*, but little is known about whether these clones are increasing or decreasing in prevalence. We therefore analyzed the MLSTs of each isolate for the presence of high-risk clone STs. Existing MLSTs were identified for 209 of the 227 isolates. The remaining 18 isolates had exact identity matches for all seven housekeeping genes but did not correspond to known MLSTs, indicating that they represent new STs. Of the 93 isolates from 1999-2002, the most common MLSTs were ST298 (8 isolates; 8.6%), ST348 (8 isolates; 8.6%), ST111 (6 isolates; 6.5%), ST639 (5 isolates; 5.4%), and ST2613 (4 isolates; 4.3%). From 2003-2009, of 101 isolates, the most common were ST639 (8 isolates; 7.9%), ST2613 (5 isolates; 4.9%), ST471 (5 isolates; 4.9%), ST298 (4 isolates; 3.9%), and ST111 (4 isolates; 3.9%). From 2017-2019, the most common were ST252 (3 isolates; 9.1%), ST646 (3 isolates; 9.1%), ST27 (2 isolates; 6.1%), and ST532 (2 isolates; 6.1%). Of these STs, two (ST111 and ST298) have been reported as high-risk clones. (ST175 and ST235, the other two well characterized high-risk clones, were not found in our cohorts). The cumulative prevalence of high-risk clones therefore decreased from 14 of 93 isolates (15%) in 1999-2002 to 1 of 33 isolates (0.30%) in 2017-2019 (Chi-Square test of linear trend, χ = 4.757, p = 0.029). These findings demonstrate that high-risk clones have been circulating at NMH but may be decreasing in prevalence.

### Presence of Aminoglycoside Modifying Enzyme Genes in P. aeruginosa Isolates

We next examined the whole genome sequences of each isolate for aminoglycoside modifying enzyme (AME) and methylase genes (Table 2). Except for *aph(3’)-IIb*, which was found in all isolates, AME genes were relatively rare. *ant(2”)-Ia* was the most common AME gene in the 1999-2002 cohort, followed by several *aadA* genes (members of the *ant(3”)* family). In the 2003-2009 cohort, the most common AME gene was *aadA6*, and *ant(2”)-Ia* was still fairly prevalent. In the 2017-2019 cohort, only genes from the *ant* family (*aadA6, aadA10, ant(2”)-Ia*) were present, perhaps due to the smaller size of this collection. None of the cohorts contained isolates with the *rmtA* or *rmtB* methylase genes. Excluding *aph(3’)-IIb*, the prevalence of isolates with at least one AME gene decreased from 16 of 93 isolates (17.2%) in the 1999-2002 cohort to 14 of 101 isolates (13.8%) in the 2003-2009 cohort to 2 of 33 isolates (6.1%) in the 2017-2019 cohort. These differences were not statistically significant (Chi-Square test of linear trend, χ = 2.292, p = 0.130).

**Table 1.**
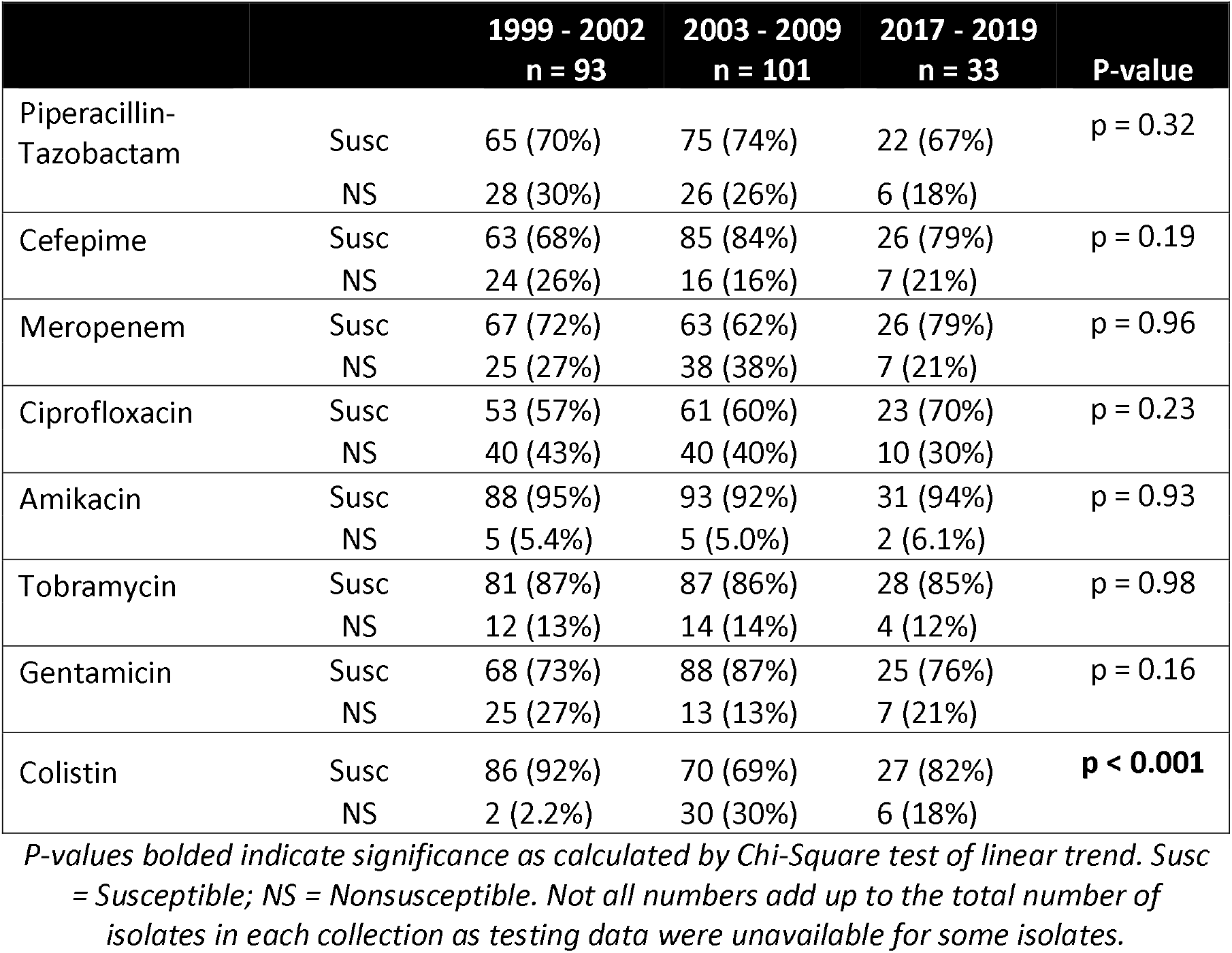
Susceptibility of *P. aeruginosa* isolates to antibiotics by time period.

**Table 2.** AME genes present in *P. aeruginosa* isolates.

| Number of Isolates with Gene (% of Isolates) |  |  |  |  |
| --- | --- | --- | --- | --- |
|  | 1999 - 2002<br>(n=93) | 2003 - 2009<br>(n=101) | 2017 - 2019<br>(n=33) | Total<br>(n=227) |
| *AAC(3)-Id* | 1 (1%) | 0 (0%) | 0 (0%) | 1 (0.4%) |
| *AAC(3)-IIIb* | 0 (0%) | 2 (2%) | 0 (0%) | 2 (0.9%) |
| *AAC(6')-Ib* | 1 (1%) | 3 (3%) | 0 (0%) | 4 (2%) |
| *AAC(6')-Ib'* | 3 (3%) | 0 (0%) | 0 (0%) | 3 (1%) |
| *AAC(6')-IIc* | 0 (0%) | 1 (1%) | 0 (0%) | 1 (0.4%) |
| *AAC(6')-II* | 1 (1%) | 0 (0%) | 0 (0%) | 1 (0.4%) |
| *aadA1* | 0 (0%) | 2 (2%) | 0 (0%) | 2 (0.9%) |
| *aadA10* | 4 (4%) | 3 (3%) | 1 (3%) | 8 (4%) |
| *aadA2* | 4 (4%) | 0 (0%) | 0 (0%) | 4 (2%) |
| *aadA6* | 4 (4%) | 5 (5%) | 1 (3%) | 10 (4%) |
| *aadA7* | 0 (0%) | 3 (3%) | 0 (0%) | 3 (1%) |
| *ant(2'')-Ia* | 8 (9%) | 4 (4%) | 1 (3%) | 13 (6%) |
| *aph(3')-Ib* | 0 (0%) | 1 (1%) | 0 (0%) | 1 (0.4%) |
| *aph(3'')-Ib* | 1 (1%) | 0 (0%) | 0 (0%) | 1 (0.4%) |
| *aph(3')-IIb* | 93 (100%) | 101 (100%) | 33 (100%) | 227 (100%) |
| *aph(6)-Id* | 1 (1%) | 0 (0%) | 0 (0%) | 1 (0.4%) |
| Isolates with at least 1 AME gene* | 16 (17%) | 14 (14%) | 2 (6%) | 32 (14%) |

\*Excluding *aph(3')-IIb*.

### Associations Between Aminoglycoside Resistance and Presence of AME Genes

We next examined whether individual AME genes as well as combinations of AME genes predicted phenotypic resistance to aminoglycosides. We focused on gentamicin and tobramycin because only four AME genes targeting amikacin were present in our collections. For gentamicin and tobramycin, we individually examined each of the AME genes but found that only *ant(2”)-Ia* showed a statistically significant association with resistance to gentamicin (OR 4.03 [1.02, 15.84], p = 0.046) or tobramycin (OR 5.06 [1.32, 19.51], p = 0.018). A statistically significant association was not found between phenotypic resistance and the presence of *at least one* AME gene with predicted activity against either gentamicin (OR 2.53 [0.88, 7.28], p = 0.084). (A similar grouped analysis was not performed for tobramycin because *ant(2”)-Ia* was the only AME gene in our collection with predicted activity against this antibiotic.) These results suggest that AME genes play a relatively minor role in causing gentamicin and tobramycin phenotypic resistance in *P. aeruginosa* at NMH.

### Genomic Characterization of an Isolate with Multiple AMEs

We next more closely examined the distribution of AME genes in our collection of isolates. As mentioned earlier, all isolates contained the *aph(3’)-IIb* gene, but most isolates did not contain additional AME genes. One exception was PS1871, an XDR isolate (nonsusceptible to piperacillin-tazobactam, cefepime, meropenem, ciprofloxacin, tobramycin, and gentamicin) that was noted to have a total of five AME genes (*aph(3’)-IIb, AAC(6’)-Ib, ant(2”)-Ia*, and two copies of *aadA1)*. PS1871 had the following MICs to aminoglycosides: 16 µg/mL for amikacin (susceptible), 16 µg/mL for tobramycin (nonsusceptible), and 16 µg/mL for gentamicin (nonsusceptible).

To further characterize PS1871, we performed long-read sequencing and combined these results with the previously obtained short-read sequencing results to obtain its complete genome. This genome was used to identify the locations of the AME genes (Figure 2). *ant(2”)-Ia* and *aadA* are located among a cluster of transposon genes, alongside *qacE* (encoding an efflux pump associated with antiseptic resistance). *AAC(6’)-Ib* and a second copy of *aadA* are located adjacent to two β-lactamase genes, *OXA-9* and *TEM-1*, and near a different copy of *qacE* as well as *dfrA1*, a trimethoprim-resistance-conferring dihydrofolate reductase. These genes are also associated with a large transposon complex. Finally, *aph(3’)-IIb* was located in a distinct region of the chromosome and was not adjacent to known AMR or transposase genes.

**Figure 2.**
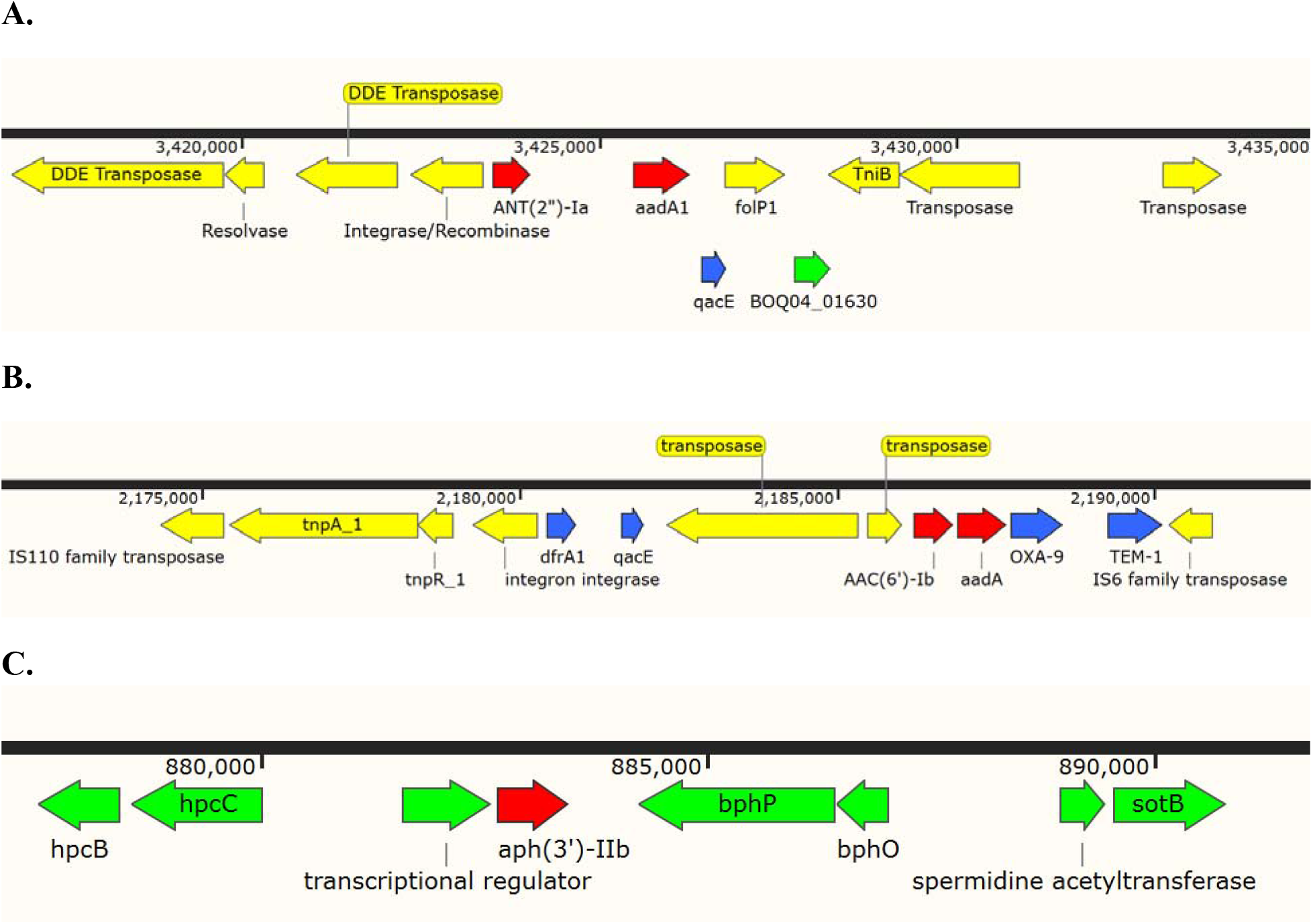
The context of AME genes present in PS1871. **A**. *ant(2”)-Ia* and *aadA1*. **B**. *AAC(6’)-Ib* and *aadA1*. **C**. *aph(3’)-IIb*. Colors represent the following: red, AME genes; blue, genes conferring resistance to antiseptics and antibiotics other than aminoglycosides; yellow, transposition elements; green, other genes.

## Discussion

In this study, we examined changes in aminoglycoside resistance rates in *P. aeruginosa* over time at our institution. We focused on bloodstream isolates and found that susceptibility rates were relatively stable from 1999 to 2019. Similar trends were observed with several other commonly used antipseudomonal antibiotics. An exception was colistin, for which nonsusceptibility rates varied substantially over time. These results should be considered in the context of several trends. An increasing prevalence of MDR and XDR *P. aeruginosa* isolates has been frequently reported, with rates of drug-resistant strains between 15% and 30% in some areas [29][30]. For example, Yayan et al. (2015) noted a significant increase in the prevalence of MDR isolates in a sample of 168 *P. aeruginosa* pneumonia isolates from 2004-2014 [31]. On the other hand, the CDC has estimated that antimicrobial stewardship efforts have led to a 29% drop in MDR *P. aeruginosa* infections in the U.S. between 2012 and 2017 [32]. Consistent with this, Abbara et al. (2019) studied the before (2007-2010) and after (2011-2014) period of an antimicrobial stewardship program intervention and found significant improvements in antibiotic usage, as well as dramatic decreases in *P. aeruginosa* resistance rates, including a drop in resistance to amikacin from 30.9% to 0.8% [33]. The antibiotic stewardship program at NMH began in the 1990s but was substantially enhanced in 2006, when it formally received funding for the administrative effort of a physician. A third potential factor influencing antibiotic resistance rates are infection control and prevention efforts. In our study, the decreased prevalence over time of high-risk clones, which are thought to be transmitted from patient to patient, could reflect the increasing success of these efforts. Antibiotic resistance is influenced by a complex set of factors, and the reasons contributing to relatively stable rates at our hospital require further study.

Our genomic analyses indicated that AME genes are relatively rare at NMH. The only exception was *aph(3’)-IIb*, which provides intrinsic resistance to kanamycin, neomycin, butirocin, and seldomycin, though not the clinically relevant aminoglycosides gentamicin, tobramycin, or amikacin [34]. In agreement with prior reports showing that *aph(3’)-IIb* is present in nearly all *P. aeruginosa* bacteria [34], we found this chromosomal aminoglycoside phosphotransferase in every isolate in our collection. Other relatively common AME genes were *ant(2”)-Ia* (5.7% of isolates), *aadA6* (4.4% of isolates), and *aadA10* (3.5% of isolates). Notably, *aadA* AMEs are members of the *ant(3”)* family of nucleotidyltransferases, which are the most common *ant* AME genes and are typically found in integrons, plasmids and transposons. These AMEs primarily confer resistance to spectinomycin and streptomycin, but not gentamicin, tobramycin, or amikacin [14]. Excluding the ubiquitous *aph(3’)-IIb*, the overall prevalence of isolates containing AME genes was 14%, less than that reported in several published studies. A French study of 120 *P. aeruginosa* bacteremia isolates collected between 1999 and 2004 showed that 25 (21%) of them contained one or more genes encoding for the AMEs *ant(2”)-I, AAC(6’)-I*, and *AAC(3)-I* [35]. In northeastern Poland, a study of 25 *P. aeruginosa* samples collected from two intensive care units between 2002-2009 detected *ant(2”)-Ia* in 36%, *AAC(6’)-Ib* in 28%, and *aph(3’)-Ib* in 8% of the isolates [36]. Holbrook & Tsodikova (2018) studied 122 MDR *P. aeruginosa* isolates from the University of Kentucky Health System and found that 41% carried *AAC(6’)-Ib*, 11% carried *AAC(3)-IV*, 6% carried *ant(2”)-Ia*, and 71% carried *aph(3’)-Ia* [37]. In Venezuela, a study of 137 *P. aeruginosa* isolates collected from clinical samples during 2010-2011 found that 49 isolates were resistant to at least one of the following aminoglycosides: gentamicin, amikacin, netilmicin, and tobramycin. Within those 49, 39 had at least one AME gene detected. The most commonly detected AME genes were *AAC(6’)-Ib, aphA1, aadB, aphA2, and AAC(3’)-IIa* [38]. There are several potential explanations for the differences between our results and these published studies. The results cited above are from different geographical locations than NMH, and AME gene prevalence may differ from region to region. In addition, the cited reports examined different types of isolates collections, including specific collections of MDR isolates, ICU isolates, or non-bloodstream isolates.

Several groups have predicted aminoglycoside resistance in bacteria other than *P. aeruginosa* based upon the presence or absence of AME genes [41]. Stoesser et al. (2013) compared phenotypes of resistance for *E. coli* and *Klebsiella pneumoniae* with genotypic predictions of resistance, and noted 96% sensitivity and 100% specificity for gentamicin [39]. Sadouki et al. (2017) compared phenotypic and genotypic resistance for ten antibiotics against *Shigella sonnei* and found only fifteen (0.45%) discordances across 3,350 isolate/antimicrobial class combinations. Only 7 of 335 isolates were resistant to gentamicin, and each of the 7 possessed either *AAC(3)-IId* or *AAC(3)-IVa*, which confer gentamicin resistance [40].

Attempts to predict aminoglycoside phenotypes from genotypes have been less successful in *P. aeruginosa* than in other bacteria. Several reports examining only one or two isolates have noted correlations. For example, de Oliveria Santos et al. (2018) reported an isolate that was phenotypically susceptible to gentamicin and resistant to amikacin, and it carried *aph(3’)-IIb* and *aph(3’)-VI*, which confer resistance to amikacin but not gentamicin [42]. Kocsis et al. (2019) examined a *P. aeruginosa* ST773 isolate and found *rmtB*, a 16S rRNA methylase, concordant with the isolate’s significant tobramycin resistance [43]. Madaha et al. (2020) looked at a pair of multidrug resistant *P. aeruginosa* strains and found that one was pan-susceptible to aminoglycosides, harboring only *aph(3’)-IIb*, while the other, which harbored *rmtB* and several other AME genes, was pan-resistant to aminoglycosides [44]. Examination of larger collections of isolates, however, have not observed these associations. Freschi et al. (2018) examined 33 *P. aeruginosa* isolates acquired from cystic fibrosis (CF) patients and seven reference genomes. They found poor correlations between genetic resistomes and phenotypic susceptibilities, noting that only for fluoroquinolones were correlations observed [45]. Jeukens et al. (2017) analyzed genetic resistomes and correlated them to resistance phenotypes in seven isolates. Only one was resistant to aminoglycosides, and this isolate harbored no identified resistance determinants apart from *aph(3’)-IIb*. These findings underscore the difficulties in linking the presence of AME genes to phenotypic aminoglycoside resistance in *P. aeruginosa*.

In this study, our ability to successfully predict aminoglycoside resistance phenotypes from the presence of AME genes was limited. This may be due to the multiple mechanisms that can account for aminoglycoside resistance in *P. aeruginosa*. In addition to AMEs and methylases, *P. aeruginosa* is capable of increased aminoglycoside efflux and impermeability [1, 46]. A limitation of our study is that these other mechanisms were not assessed. Likewise, we did not measure expression of AME genes, and it is possible that some of these genes were “silent.” Broader experimental and genomic studies will be necessary to better predict aminoglycoside phenotypes from *P. aeruginosa* genomes.

We examined in detail the genome of one XDR isolate, PS1871. We found that of the five AME genes present in this isolate, four occurred in pairs, and each pair was found in the context of multiple transposon genes. Additionally, both pairs were near a *qacE* efflux pump gene, and one pair was located next to β-lactamase genes *OXA-9* and *TEM-1*. These findings suggest that these four AME genes are capable of being transferred by horizontal transfer of genetic elements and that this process may be driven by exposure to antibiotics and disinfectants.

Our findings suggest that resistance to aminoglycosides remains relatively stable and uncommon at our hospital and is largely driven by mechanisms other than AME genes. However, XDR strains such as PS1871 may harbor multiple AME genes, and ongoing vigilance is required to prevent their spread.

## Data Availability

All data used to generate the tables and figures for this manuscript are available upon request.

## ACKNOWLEDGEMENTS

The project was funded by the National Institutes of Health: R01AI118257, U19AI135964, K24AI04831, and R21AI129167 (all to A.R.H.).

